# Estimating COVID-19-induced Excess Mortality in Lombardy

**DOI:** 10.1101/2021.11.17.21266455

**Authors:** Antonello Maruotti, Giovanna Jona-Lasinio, Fabio Divino, Gianfranco Lovison, Massimo Ciccozzi, Alessio Farcomeni

## Abstract

We compare the expected all-cause mortality with the observed one for different age classes during the pandemic in Lombardy, which was the epicenter of the epidemic in Italy and still is the region most affected by the pandemic. A generalized linear mixed model is introduced to model weekly mortality from 2011 to 2019, taking into account seasonal patterns and year-specific trends. Based on the 2019 year-specific conditional best linear unbiased predictions, a significant excess of mortality is estimated in 2020, leading to approximately 35000 more deaths than expected, mainly arising during the first wave. In 2021, instead, the excess mortality is not significantly different from zero, for the 85+ and 15-64 age classes, and significant reductions with respect to the 2020 estimated excess mortality are estimated for other age classes.

## 1 Introduction

Incidence indicators based on official reporting underestimate the true number of cases since there exists a vast proportion of asymptomatic or mildly symptomatic patients, among all infected individuals, who are not detected [5, 15, 14, 16]. Complex methods are then required to provide reasonable forecasts of the epidemic evolution [2, 11, 1, 8] and avoid unreliable predictions, which make people worry (on this point, see e.g. [7, 9]). The attention has recently moved to the analysis of mortality, as the gold standard measure of the impact of COVID-19 worldwide [3, 10]. This is mainly because mortality data suffer less of underreporting, measurement errors and other data issues, i.e. are in general more reliable. We would like to contribute to this growing literature by introducing a generalized linear mixed model to estimate COVID-19-induced excess mortality in Lombardy, Italy.

Italy was dramatically hit by the COVID-19 pandemic, with 4.8 million cases and 132000 deaths as of October 31st, 2021. Lombardy, the most populated and industrialized Italian region, was the epicenter of the outbreak, in particular during the first wave in early 2020. It experienced one of highest case-fatality rate with nearly 900.000 cases (about one fifth of those registered in Italy) and nearly 34.000 deaths out of 10 million inhabitants (for a discussion see e.g. [13]). The impact of COVID-19 on mortality rates is rather clear and can be easily depicted by a simple data visualization. Quantifying the excess deaths induced by COVID-19 is more difficult and may reveal both direct and indirect effects of the pandemic on mortality. Statistical methods have been recently introduced ranging from simple averaging methods [12] to structured models [6, 17, 4].

Here we introduce a model to estimate excess mortality in 2020 and 2021 in Lombardy stratified by various age classes. The proposed generalized linear mixed model, in the spirit of [18], is able to capture year-specific trends, seasonality and overdispersion in the number of all-cause deaths and is based on historical data collected on a weekly basis from 2011 to 2019.

## 2 Data and Methods

Open-source daily all-cause Italian mortality data, stratified by Region and age classes, from 2011 to 2021 are available from the Italian National Institute of Statistics (ISTAT). These data were downloaded on October 25th 2021 and temporally aggregated at the week level. The weeks number 53, present in some years, were dropped from the analysis. Data on historical all-cause mortality are displayed in Figure 1. Seasonality is clearly present, and is shown for all the considered years. Peaks are rather evident at the major COVID-19 waves, with a maximum of 7729 weekly deaths at the end of March 2020, during the first pandemic wave. Similar behaviors, on different scales, are observed for age-specific data, not reported here for the sake of brevity.

**Figure 1:**
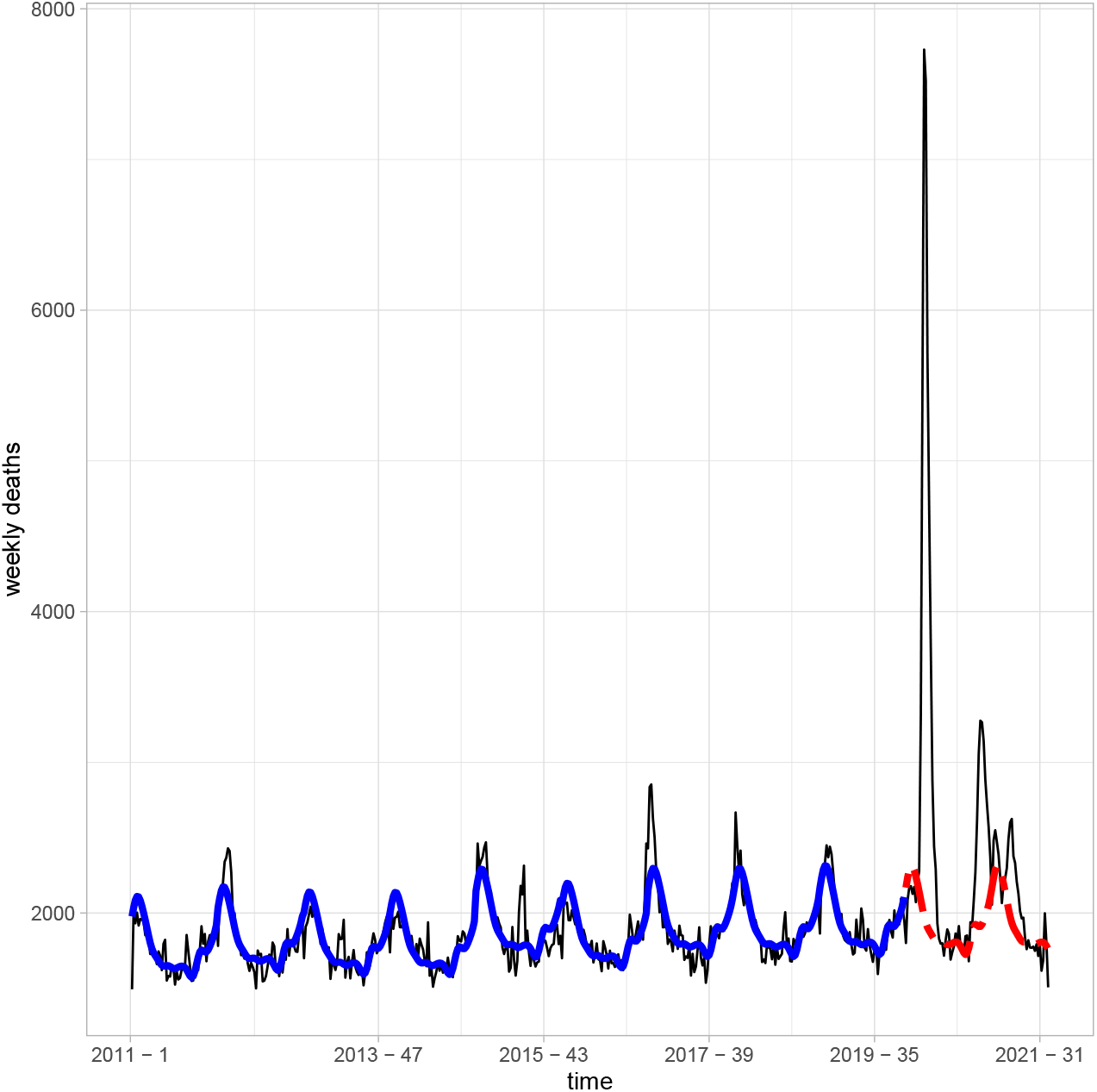
Observed total number of deaths per week (solid black line) and model’s estimated values (thick solid line). The dashed (red) line shows the expected weekly deaths under the historical model for years 2020 and 2021.

The crucial aspect in estimating excess mortality is the definition of reliable benchmark mortality model, i.e. a model providing *good* estimation of the expected mortality under pre-epidemic conditions. We model the weekly mortality *Y*_*tj*_ at week *t* = 1, …, 52 and years *j* = 2011, …, 2019. The count data process on weekly deaths is modeled with a Negative Binomial distribution, i.e.

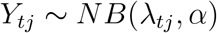

with

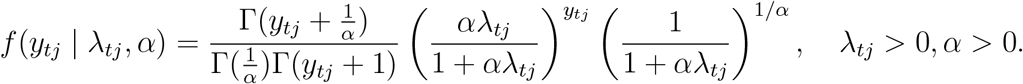

In count data analysis, the interest is usually focused upon the parameter vector *λ* = (*λ*_1,2011_, …, *λ*_*tj*_, …, *λ*_52,2019_) which is modelled, in a regression context, by defining a generalized linear mixed model with log link for the analysed response, i.e.

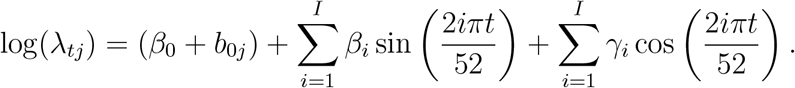

Cyclical patterns, with-year fluctuations, are captured by Fourier series, whose term *I* is defined by using model selection criteria; to account for year-specific mortality baselines and to accomodate, at least partially, autocorrelation of the weekly counts, random effects *b*_0*j*_ *∼ N* (0,σ) are introduced. Model’s fitting to the data is shown in Figure 1 (see the thick solid line). The weekly predictions of mortality data for years 2020 and 2021 are based on the 2019 year-specific conditional best linear unbiased predictions of the generalized linear mixed model. A simplified assessment the weekly excess mortality results from subtracting the predicted mortality from the observed weekly mortality under the pandemic. Finally, the weekly estimates of excess mortality are summed over the year (or part of it, for 2021).

## 3 Results and Discussion

It is well known that mortality rates are age-specific. Accordingly, different models are fitted to observed deaths by age classes. Five age classes are considered here, namely (0-14), (14-64), (65-74), (75-84), 85+. This is very useful to detect if and how COVID-19 has an impact on mortality at different age classes of the population and to plan interventions to protect at risk sub-populations.

Results are shown in Table 1. Prediction intervals are found through parametric bootstrap; if zero is included in the intervals, no difference with the expected number of deaths is estimated, i.e. there is no excess mortality. The first interesting result is that no excess mortality is estimated neither in 2020 nor in the first 34 weeks of 2021 for the youngest sub-population aged (0-14); indeed, a significant reduction in the number of deaths is estimated for this specific subpopulation in 2021. This reinforces the idea the children have very low risks of complications induced by COVID-19 infections. For all the other classes, significant excesses are estimated in 2020, leading to approximately 35000 more deaths than expected. The largest excess of deaths, in relative terms, is in the classes 65-74 and 75-84, with an excess of approximately 28%. A 25% excess is estimated in the oldest class 85+. For the working age class 15-64, instead, the estimated excess mortality is approximately 18%. There is, thus, a clear age effect on the risk of death induced by COVID-19. As a by-product of the analysis, we are able to detect weeks where the estimated excess mortality is relevant. In Lombardy, a large part of the excess of mortality is estimated during the first wave on March-April 2020, though similar behaviors, with smaller effects, are estimated in Autumn 2020.

**Table 1:**
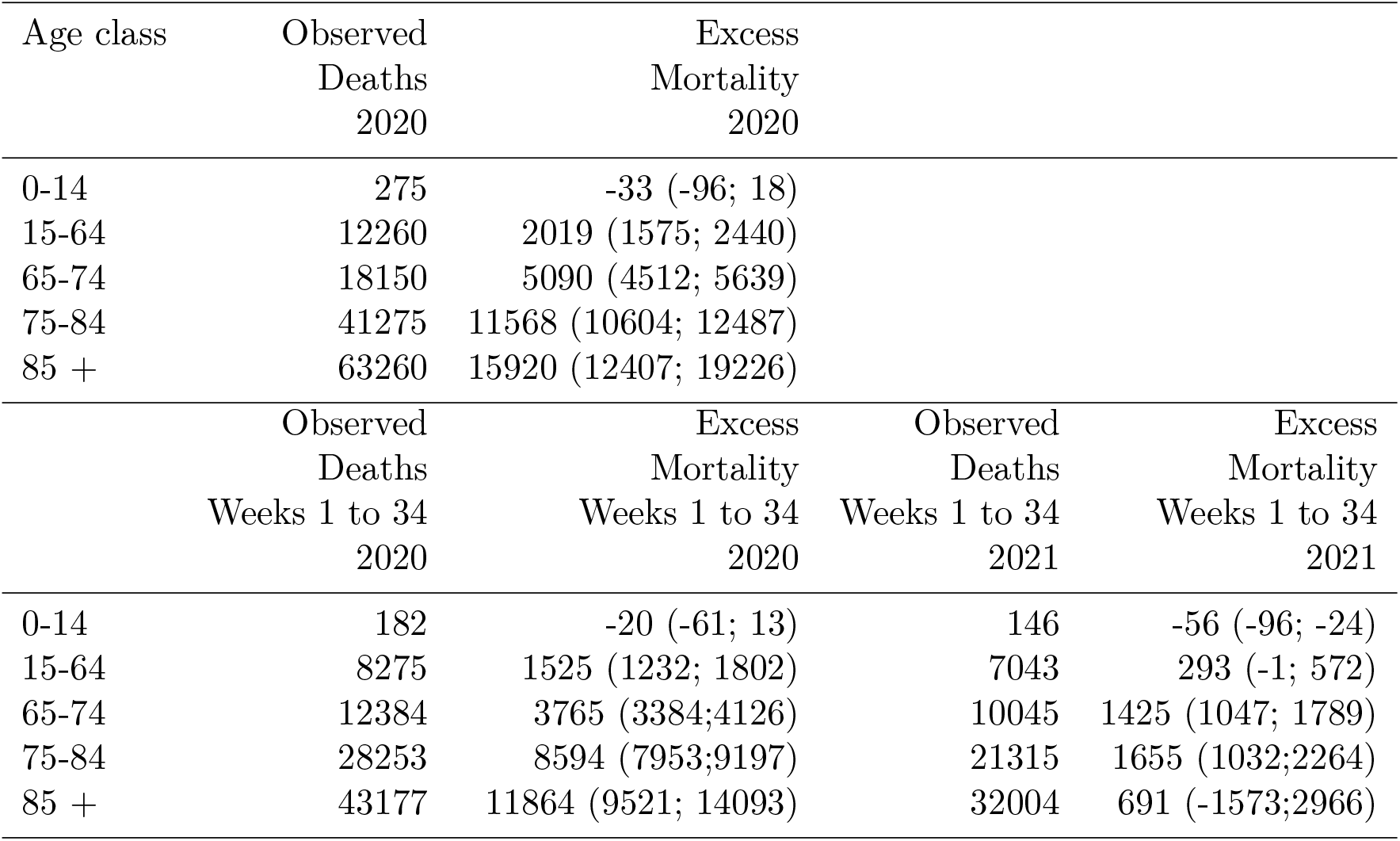
Estimated excess mortality, with 95% prediction intervals in brackets.

According to our analysis, the population 65+ has higher risks of death induced by the COVID-19 epidemic. This justifies the priority assigned to the elderly during the vaccination process, as they showed the highest mortality both in absolute and relative terms. As we do not have data for the entire 2021, we focus on the first 34 weeks (up to the end of August), for which we have complete data. Excess mortality is not significantly different from zero for the 85+ and 15-64 age classes, and the estimated excess mortality shows significant reductions with respect to 2020 estimated values for other age classes. The 65-74 and 75-84 classes are the ones more at risk, with 14.2% and 7.8% of excess mortality estimated; this is somehow not surprising because the number of not vaccinated people is still quite relevant in these age classes, according to the official statistics, and these subpopulations were the latest to be vaccinated among the elderly. The positive effect on excess mortality reduction of the vaccine can be better appreciated if we compare excess mortality in the first 34 weeks of 2020 and 2021. A strong reduction in the excess mortality induced by COVID-19 is estimated, even in age classes showing a significant excess. Estimates clear show that without the vaccine, the excess mortality is relevant in all age classes. In 2021, instead, the vaccine limits both the circulation of the virus and, more importantly, the rise of complications possibly leading to death. This results is even more robust as non-pharmaceutical interventions, as e.g. lockdowns, were quite light during 2021 and much heavier in 2020.

## Data Availability

All data produced in the present study are available upon reasonable request to the authors

## Notes

### Competing Interest Statement

The authors have declared no competing interest.

### Funding Statement

The research has been partially supported by the Ministero dell'Istruzione,
dell'Universita' e della Ricerca, project number FISR2020IP\_00156
"Modelli statistici inferenziali per governare l'epidemia" (SMIGE).

